# Combining Clinical Embeddings with Multi-Omic Features for Improved Patient Classification and Interpretability in Parkinson’s Disease

**DOI:** 10.1101/2025.01.17.25320664

**Authors:** Chaeeun Lee, Barry Ryan, Riccardo E. Marioni, Pasquale Minervini, T. Ian Simpson

## Abstract

This study demonstrates the integration of Large Language Model (LLM)-derived clinical text embeddings from the Movement Disorder Society Unified Parkinson’s Disease Rating Scale (MDS-UPDRS) questionnaire with molecular genomics data to enhance patient classification and interpretability in Parkinson’s disease (PD). By combining genomic modalities encoded using an interpretable biological architecture with a patient similarity network constructed from clinical text embeddings, our approach leverages both clinical and genomic information to provide a robust, interpretable model for disease classification and molecular insights. We benchmarked our approach using the baseline time point from the Parkinson’s Progression Markers Initiative (PPMI) dataset, identifying the Llama-3.2-1B text embedding model on Part III of the MDS-UPDRS as most informative. We further validated the framework at years 1, 2, 3 post baseline, achieving significance in identifying PD associated genes from a random null set by year 2 and replicating the association of MAPK with PD in a heterogenous cohort. Our findings demonstrate that the combination of clinical text embeddings with genomic features is critical for classification and interpretation. LLM text embeddings not only increase classification accuracy but also enable interpretable genomic analysis, revealing molecular signatures associated with PD progression.

## Introduction

Parkinson’s disease (PD) is a neurodegenerative disorder resulting from the death of dopamine producing cells in the substantia nigra. It is one of the most common neurodegenerative disorders and affects over 6 million individuals worldwide (Bloem et al., 2021). There are many known associations with the development of PD. For example, individual point mutations in genes such as *LRRK2* and *SNCA* can cause monogenic forms of the disease; however, their penetrance is low. Further, some cases of PD have been associated with environmental factors; most commonly exposure to toxins (Klein and Westenberger, 2012).

Overall, PD is a disease which poses many challenges to researchers due to heterogeneity among patients presentations, symptoms and more. The development of PD via researched genetic alterations plays a role in up to 10% of cases (Klein and Westenberger, 2012). For the majority, patients are labelled as idiopathic, meaning no cause can be found for the onset of disease. Once diagnosed, the trajectory and symptoms of patients are unknown. While most may associate the disease with motor symptoms of freezing, tremors and gait, non-motor symptoms such as cognitive decline and smell disorders can be equally debilitating. Moreover, not every individual with PD will experience the full spectrum of PD symptoms (Bloem et al., 2021). In most cases, the reverse is true, where patients will only experience a subset of symptoms, but which symptoms, to what degree and how quickly all remain unanswered questions.

As a result, great efforts have been undertaken to identify, quantify and track the motor and non-motor symptoms of PD patients. The best globally accepted metric for doing so is the Movement Disorder Society Unified Parkinson’s Disease Rating Scale (MDS-UPDRS) (Goetz et al., 2008). The MDS-UPDRS examination is a four part examination consisting of : Part I – Self assessment examination by patient or carer, Part II - Examination of non-motor symptoms by clinician, Part III - Examination of motor symptoms by clinician, Part IV - Examination of motor complications by clinician. The MDS-UPDRS has been established as a clinically insightful tool for monitoring and tracking the symptoms of PD patients. Brumm et al. (2023) used large parts of the MDS-UPDRS to identify clinically meaningful milestones which can be used to track the trajectory of patients. Skorvanek et al. (2015) were able to identify a relationship between the non-motor items of MDS-UPDRS and the quality of life in PD patients.

Previous research has identified genetic and biological mechanisms that can be significantly associated with a change in MDS-UPDRS score. Davis et al. (2016) found that alterations in the *GBA* gene were associated with progression in part 3 of the MDS-UPDRS in 733 PD patients. Li et al. (2018) were able to associate patterns of grey matter intensity in the putamen with a decrease in MDS-UPDRS part 3 scores early in the disease course of 392 patients. Despite promising findings, rarely do these studies take into account the heterogeneity of patient symptoms when attempting to make such associations with PD.

To compensate for variability, previous studies have either focused on identifying key phenotypic variables and their importance or looked at splitting patients into subtypes. Das et al. (2024) utilised patient questionnaire data to improve classification accuracy for PD. Their approach integrated motor and non-motor features, employing feature selection methods to reduce redundancy and highlight significant features with the best predictive power. Fereshtehnejad et al. (2017) used cluster analysis to group PD patients into three subtypes: mild motorpredominant, intermediate, and diffuse malignant. While clustering identified key features deemed most relevant to the subtype classification, the study developed a categorical subtype definition to apply results at an individual level based on critical features identified through Principal Component Analysis (PCA).

Since subtyping systems are often used for their prognostic capabilities (Ygland Rödström and Puschmann, 2021), it is important that this variability is accounted for without carrying over biases learned from specific cohorts when performing inferences. Our approach bypasses identification of the best classifier variables by leveraging LLM embeddings, encoding entire questionnaire responses into a high-dimensional latent space. This method captures nuanced relationships within the data without presupposing feature importance, avoiding cohort-specific biases and ensuring adaptability to new datasets.

LLM-based embeddings have recently begun to surpass encoder only models such as BERT (Devlin et al., 2019) and Sentence-BERT (Reimers and Gurevych, 2019) in many tasks, as demonstrated on the Massive Text Embedding Benchmark (Muennighoff et al., 2023; Wang et al., 2024; Lee et al., 2025). They eliminate the need for curated hard negative sets or highly diverse datasets, which are unsuitable for semi-structured questionnaire data composed of repeated question texts and predefined response options. LLMs also offer significantly longer context windows, enabling them to encode entire questionnaire sections, which is often infeasible with smaller encoder-only text embedding models.

Further, our method uses patient networks, which have shown promise in a number of areas for capturing inter patient variability. Such a network represents patients as nodes and identifies relationships between patients with similar, molecular, genetic, or clinical phenotypes. Li et al. (2022) use patient similarity to improve cancer subtype predictions. Similarly, Zhang et al. (2022) show that stratification of liver cancer subtypes is improved when integrating genomic modalities using a patient similarity framework. Deriving patient relationships from text embeddings encoding disease symptoms is a novel approach for overcoming patient heterogeneity in PD.

The focus of this paper is to use the MDS-UPDRS examination to form a patient similarity network which captures the inter-patient variability in symptoms and to identify informative genes and molecular mechanisms using a biologically interpretable Graph Neural Network (GNN) model. We use text embedding models to capture the full spectrum and context of the MDS-UPDRS. We show that capturing this context improves on similarity measures calculated from feature selection or subtyping. We highlight the importance of generating patient similarity networks from MDS-UPDRS embeddings and we show that combining these embeddings with multiple genomic measures can yield insights into disease mechanisms. We perform the primary analysis using the baseline time point of the Parkinson’s Progression Markers Initiative (PPMI) dataset and validate at subsequent time points of 1, 2 and 3 years post diagnosis.

## Methods

### Datasets

Data was obtained from the publicly available PPMI resource (Marek et al., 2018). The PPMI dataset tracks, longitudinally, the progress of PD patients in several genomic modalities and in the MDS-UPDRS. Three genomic modalities, namely Messenger RNA expression (mRNA), Cerebral Spinal Fluid Proteins (CSF) and DNA methylation (DNAm) were included in the analysis. The genomic data from PPMI, with the exception of CSF, was generated from blood samples. These modalities were selected due to their diverse coverage and relevance to the disease (Redenšek et al., 2018; Wüllner et al., 2016). The patient data availability in each modality per time point is shown in supplementary table 1.

We included parts 2 and 3 of the MDS-UPDRS only. We selected these parts as they cover a range of motor and non-motor symptoms and should be more standardised across participants as they are conducted by a clinician. In contrast, part 1 is a self-assessment, and it has been shown to be less reliable than the other parts (Evers et al., 2019). Part 4 is only conducted on a subset of PD patients if they have a motor complication, thus there was too much missingness to support its inclusion.

For the preliminary analysis, we focused on the baseline time point. This time point was used to identify the best text embedding model, pooling strategy, model hyperparameters and section integration strategies. In PPMI, PD patients recruited at baseline had received their diagnosis less than two years previously, had not taken any PD medication, and had developed at least one motor symptom (Marek et al., 2018). Given these criteria, at baseline we would expect the MDS-UPDRS to capture differences between PD patients and Healthy Control (HC), however, as of writing, there are no known genomic biomarkers for PD which can be measured from the blood. Thus, our research investigates if using patient similarities based on text embeddings can be used as a medium to identify genomic signatures at baseline. We further extended this analysis to subsequent yearly time points. The hypothesis is that it is more likely patients at later time points will have a stronger disease signature, which will be reflected in their blood and better captured by the genomic modalities.

### Generating Clinical Embeddings from MDS-UPDRS

To generate clinical phenotype embeddings from the MDS-UPDRS responses, we employed LLMs to encode responses from the questionnaire into dense vector representations. An autoregressive LLM processes input sequences *X* = *{x*1, *x*2, …, *xn}* sequentially, generating a hidden state *hi* for each token *xi* based on the tokens preceding it: *hi* = *f*_*θ*_ (*x*1, *x*2, …, *xi*; *M*), where *f*_*θ*_ denotes the model’s transformation function parameterized by *θ*, and *M* represents the attention mask. The attention mask ensures that token *xi* only attends to tokens *{x*1, *x*2, …, *xi}*, preserving the autoregressive property of the model. This sequential processing allows each hidden state *hi* to capture cumulative contextual information up to the *i*-th token.

### Pooling Strategy

As LLM embeddings are derived from the latent vectors of the final layer corresponding to each input token, a pooling strategy is required to combine these vectors into a unified representation of the input sequence. Mean pooling averages the last layer hidden states of all tokens in the sequence. To account for variable sequence lengths and padding tokens, we incorporate the attention mask *M*, which assigns a binary value *mi* to each token, where *mi* = 1 if the token is valid and *mi* = 0 otherwise. The mean-pooled embedding is computed as:

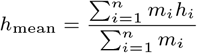

This approach ensures that only valid tokens contribute to the final representation, aggregating information across the entire sequence. However, this method can dilute critical contextual signals, especially for longer sequences.

Last token pooling directly uses the embedding of the final valid token in the sequence, as determined by the attention mask. *k* represent the index of the last valid token, where *k* = max *{i* | *mi* = 1*}* and the last token embedding is then

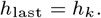

This strategy leverages the autoregressive nature of the model, where the last token embedding captures the model’s understanding of the entire input sequence, providing a concise and comprehensive representation.

### MDS-UPDRS Embedding Generation and Models

Input sequences were constructed from the MDS-UPDRS questionnaire by combining the full text of questions, associated answers, and any relevant instructions for the patient or examiner. For each patient, responses from Sections 2 and 3 were formatted as question-answer pairs, ensuring the semantic relationship between questions and answers was preserved (Supplementary Figure 5). We evaluated three configurations: embedding Section 2 or Section 3 individually and jointly embedding both sections as a single input sequence. For joint embeddings, all questions and answers from both sections were concatenated in order.

We utilised a range of autoregressive LLMs for generating embeddings from the MDS-UPDRS questionnaire, including Llama-3.2-1B, Llama-3.1-8B and Mistral-7B-v0.1. These models were chosen to explore the effects of varying parameter sizes and embedding dimensions on performance. Additionally, both base models and instruction-tuned variants were evaluated to assess how task-specific fine-tuning influences the quality of the generated embeddings.

We used a maximum sequence length of 4024 tokens to ensure the selected sections of the questionnaire could be processed without truncation. Temperature and sampling parameters were disabled during embedding extraction to ensure deterministic outputs. These settings allowed us to isolate the impact of model architecture and size on embedding quality without introducing variability from generation-specific parameters.

### Combining Embeddings and Genomics via Patient Similarity

Patient networks were generated from the MDS-UPDRS text embeddings by calculating the Pearson similarity between patients in the embedding space. The network was generated using the K-nearest neighbours algorithm, whereby an edge was generated between each patient and their 15 closest neighbours in this space. For comparison, logistic regression was also performed on the MDS-UPDRS assessments for feature selection. The selected features were used to calculate the Pearson correlation between patients, and a second patient similarity network was generated via the same KNN algorithm with K = 15.

A GNN framework was used to combine the patient similarity network with genomic modalities using the Multi-Omic Graph Diagnosis (MOGDx) architecture (Ryan et al., 2024). GNN models are well suited to this problem as they provide the ability to learn from the network structure as well as the node features. MOGDx is a framework which takes as inputs patient networks and any number of genomics and trains a specified GNN model. MOGDx uses an encoder architecture for dimensionality reduction of the genomic modalities and combines them via mean pooling during the training phase of the GNN. The latent mean pooled vectors are then provided to the GNN as node features. The GNN and the encoder are trained via a shared loss function, ensuring the model is learning both the structure of the patient network and the genomic inputs.

The encoder architecture, shown in Figure 1 Panel C, is an interpretable biological architecture, coined PNet, and, is described in the paper by Elmarakeby et al., 2021. PNet consists of a multi-layered hierarchical network curated from the Reactome database (Elmarakeby et al., 2021). A fully connected linear layer connects the genomic inputs to a user-defined list of genes. The list of genes used were obtained from the Disgenet database (Piñero et al., 2020). Disease codes C0030567, C0242422, and C0947810 cover known PD disease associations. The negative gene set was generated by randomly selecting genes from the gene layer of the Reactome network which were not previously included in the positive PD set. This full gene list, provided in the supplementary, connects to low level pathways as defined by the Reactome database. Subsequent network layers connect lower level pathways to higher level pathways, creating the hierarchical structure.

**Fig. 1:**
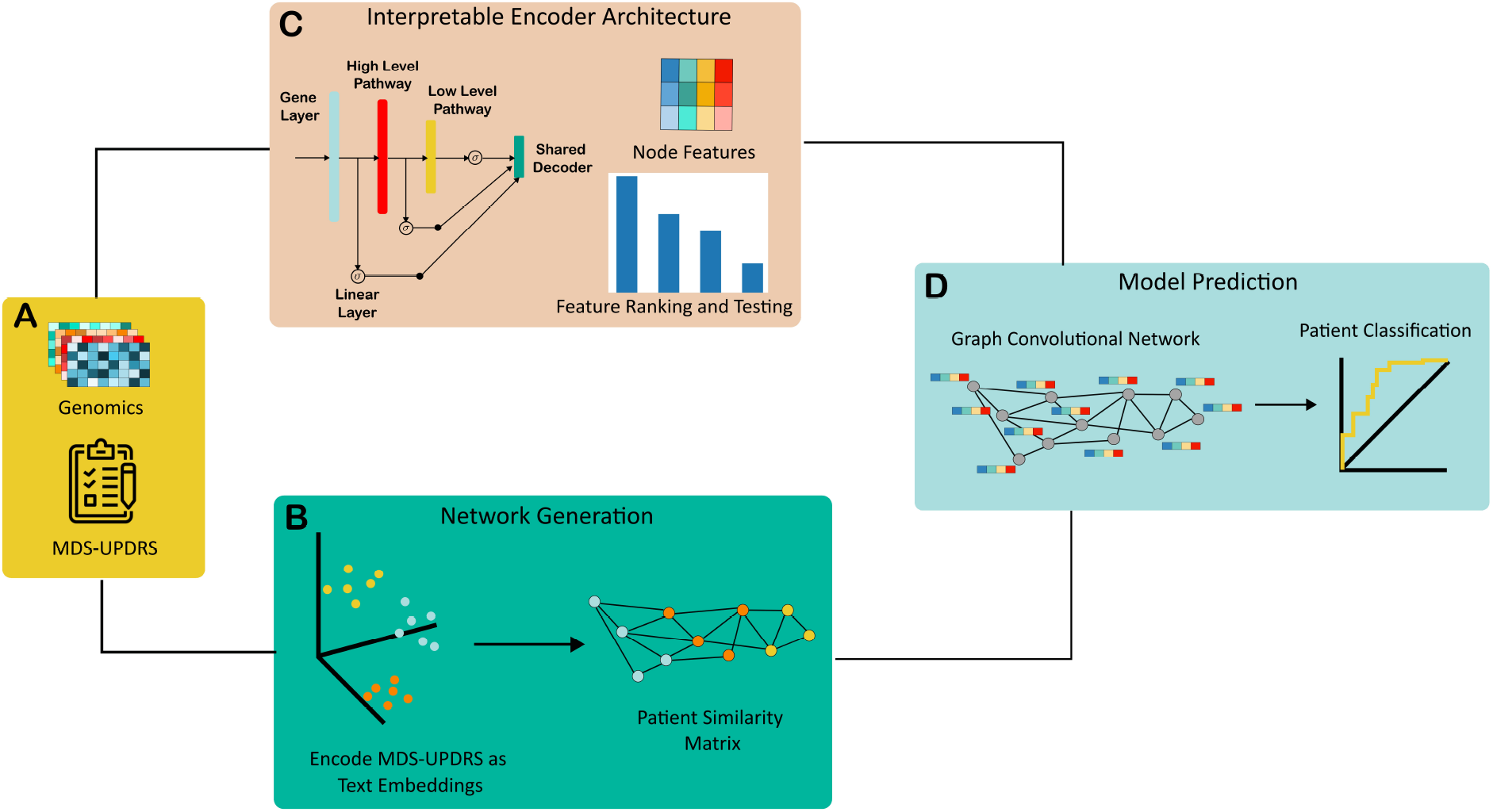
Overview of methodology pipeline. Genomic and MDS-UPDRS input data types are shown in panel A. Panel B shows the generation of patient similarity networks from text embeddings. Panel C shows a simplified layout of the PNet architecture and a sample feature importance ranking. Panel D, shows the combination of patient network with the PNet encoder architecture for patient classification.

### Classification and Feature Importance

Classification was performed between PD patients and HC. Performance was compared between models with similarity networks generated from MDS-UPDRS and directly from genomic modalities. For the networks generated from MDS-UPDRS, performance was compared with and without text embeddings, between different pooling strategies and text embedding models described previously. Performance was compared by generating standard errors across five-fold cross validation splits. For the best performing models, performance was further compared between models abilities to identify statistically significant informative features. Testing the classification performance of a model which learns from MDS-UPDRS network structure alone was not conducted, as this model would not have any meaningful molecular inputs into the interpretable encoder architecture.

Feature importance was conducted by identifying the attributions of the input features and the intermediate PNet layers using the layer conductance algorithm (Dhamdhere et al., 2018). This algorithm performs stepped feature ablation between two input vectors to attribute the effect of each feature or node in a neural network to the prediction. As our encoder network consists of known biological interactions and each layer has biological meaning, we can use this to identify the most informative genes and pathways. The mean absolute distance of each nodes’ attribution at each layer for each cross validation split was calculated and compared to a chi squared background distribution to find the features which had a statistically significant (p-value *<* 0.01 - bonferroni corrected for cross validation splits) attribution greater than zero. Further, the gene set enrichment algorithm was modified to compare the variation in attribution of the PD gene set to a randomly generated null gene set. The expectation is that genes with larger variations, i.e. positive contributions to one class and negative to the other, are the most informative. The variation in gene attribution was standardised and aggregated across cross validation splits and modalities and tested for significance (p-value *<* 0.05). We averaged this test across cross-validation splits to identify reliable genes which have positive associations with PD in all splits. Thus, there is a single hypothesis being tested, that genes with a larger variation will be from the PD set, therefore this test does not require multiple correction.

## Results

We evaluated patient classification performance at four time points defined in the PPMI study protocol. Each experiment classifies HC from PD patients. We used the baseline time point to establish the optimal configuration for text embedding model selection, MDS-UPDRS section inclusion, pooling strategy and model hyperparameter selection. We further tested the optimal configuration at subsequent time points to assess the molecular features identified at each time point. We validated the identified features by performing a gene set enrichment test to see if our model significantly identified PD associated genes from a random null set. We compared each model to a model which used logistic regression for feature selection, from which similarity between patients was measured instead of text embeddings. We also compared to a model which did not use the MDS-UPDRS assessment to derive patient similarity, instead measuring similarity directly from the genomic inputs. We did not train a model based on MDS-UPDRS derived patient similarity only, as such a model would not have any meaningful molecular inputs.

### Classification Results

Table 1 presents a comparison of the best performing classification results at baseline using our method alongside two ablation settings: one using only genomics data without MDS-UPDRS questionnaire data, and another employing logistic regression on the questionnaire data to build patient similarity networks. Genomics-only setting consistently underperformed compared to methods that included clinical patient networks, which consistently improved accuracy by over 20%. This improvement underscores the diagnostic utility of incorporating clinical patient networks alongside molecular data. Logistic regression achieved a mean accuracy of 0.972 (*±*0.013), suggesting that the patient cohort exhibited strong symptoms of PD at baseline. This is supported by the PPMI study’s recruitment criteria, which require participants to have been diagnosed with PD for up to two years, and have developed at least one PD associated motor symptom. While these criteria aim to include relatively early-stage patients, this approach means that participants at baseline show pronounced indicators of PD.

**Table 1.** Results of different model configurations for patient similarity network generation at baseline. Graph Convolutional Network (GCN) was deployed as the GNN for each model. Either text embedding models such as `Llama-3.2-1B` or `Mistral-7B-v0.1` were used to derive MDS-UPDRS text embeddings from which similarity was calculated. Similarity was also calculated from Logistic Regression and Genomic features, coined Genomics Only. The configurations resulting in the highest accuracy and F1 score are presented.

| Text Embedding Model | MDS-UPDRS Section | Pooling Strategy | GNN Model | Hidden Layer | Acc. | F1 |
| --- | --- | --- | --- | --- | --- | --- |
| Llama-3.2-1B | 3 | last token | GCN | 16 | $0.995 \pm 0.0027$ | $0.994 \pm 0.0037$ |
| Llama-3.2-1B-Instruct | 2 & 3 | last token | GCN | 32 | $0.995 \pm 0.0027$ | $0.994 \pm 0.0037$ |
| Llama-3.1-8B | 2 & 3 | last token | GCN | 64 | $0.995 \pm 0.0027$ | $0.994 \pm 0.0037$ |
| Llama-3.1-8B-Instruct | 2 & 3 | last token | GCN | 16 | $0.995 \pm 0.005$ | $0.994 \pm 0.007$ |
| Mistral-7B-v0.1 | 2 & 3 | last token | GCN | 16 | $0.995 \pm 0.0027$ | $0.994 \pm 0.0037$ |
| Mistral-7B-Instruct-v0.1 | 3 | last token | GCN | 64 | $0.995 \pm 0.0027$ | $0.994 \pm 0.0037$ |
| Logistic Regression | 3 | N/A | GCN | 16 | $0.972 \pm 0.013$ | $0.969 \pm 0.014$ |
| Genomics Only | N/A | N/A | GCN | 64 | $0.768 \pm 0.036$ | $0.762 \pm 0.053$ |

Integrating patient similarity network constructed using LLM text embeddings have demonstrated best performance, with a mean accuracy of 0.995 (*±*0.0027) for the best model and achieving accuracy of 1 in many instances. This suggests a promising approach for providing nuanced phenotype representations, which can identify subtle disease manifestations that traditional linear classifiers, such as logistic regression, may overlook. Paired t-tests (supplementary figure 3) show that these models do significantly outperform the logistic regression counterpart, motivating their inclusion. There was no significant difference between text embedding models, highlighting all of their capability to identify meaningful similarities between the symptoms of PD patients (supplementary figure 1 and table 2).

We evaluated four different configurations for questionnaire embeddings: embedding sections 2 and 3 of the MDS-UPDRS questionnaire separately and jointly as a single input. The best performance was achieved by jointly embedding sections 2 and 3 as a single input, or from using only section 3. This result suggests that the symptoms captured by section 3 of the MDS-UPDRS are the most meaningful for classifying PD patients from HC. This section assesses the motor signs of PD, requires detailed motor examination and includes direct questions on the Hoehn and Yahr stage, thus, this finding is to be expected.

Since LLM embeddings are obtained from the latent vectors of the last layer associated with each input token, a pooling strategy is needed to aggregate these vectors into a single comprehensive representation of the input sequence. Comparing two different embedding strategies—mean pooling and last token pooling—across various models, last token pooling surpassed mean pooling in performance consistently across all models, reaching up to 0.995 (*±*0.0027) mean accuracy (supplementary figure 2).

### Interpretability

We used the Llama-3.2-1B text embedding model, with patient similarity derived from section 3 of the MDS-UPDRS, last token pooling strategy and a Graph Convolutional Network (GCN) hidden dimension of 32 for our validation experiments at subsequent time points. In figure 2, it can be seen that the baseline model does not statistically significantly identify the PD genes from the null gene set, despite almost perfect classification performance. The reason for this is due to the strong predictive power of the network structure compared to the relatively weak genomic predictive power. This model does identify disease associated genes, such as *MAPK1*, as per table 2, however, it does not rank a sufficient number of these genes highly to reach significance.

**Fig. 2:**
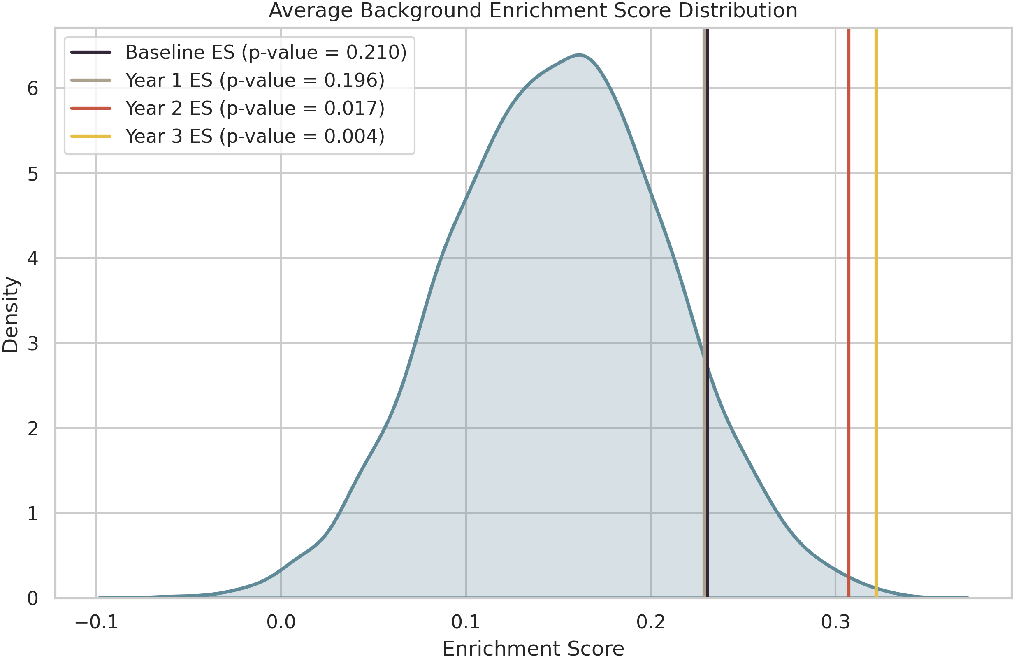
Enrichment Score (ES) for optimal model configuration at each time point. The background distribution presented was estimated based off the individual distributions of each time point. We see that both the ES increases and p-value significance decreases with time, mirroring the progressive nature of PD.

**Table 2.**
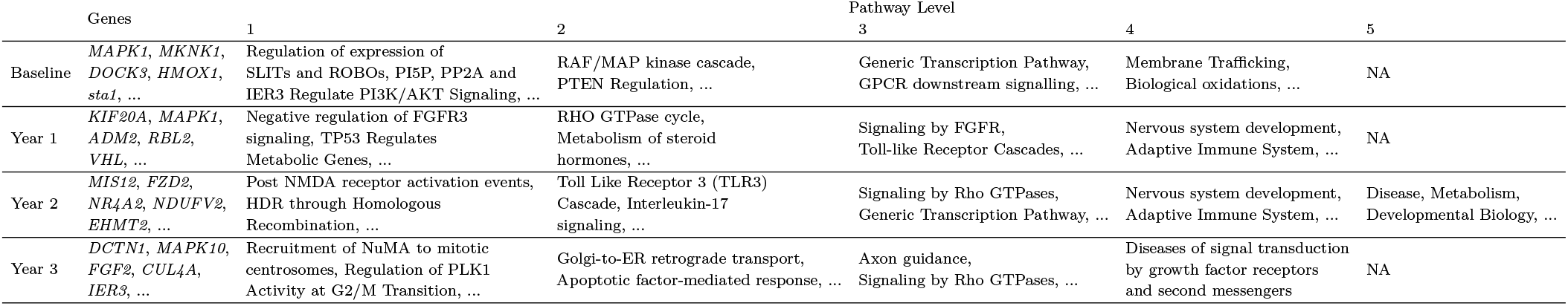
Intermediate neural network features which had a statistically significant (*p* * *value <* 0.01) non-zero attribution to the prediction across all cross-validation splits. For a complete list, along with p-values, see supplementary file 4.

Conversely, by year 2, there is sufficiently strong disease signature in the combined genomics of mRNA, CSF and DNAm to reach statistical significance. Both models, trained at years 2 and 3, rank PD genes highly, with the model trained at year 3 achieving the highest ES and lowest p-value. This result makes sense in the context of the disease. PD is a progressive disorder in which patients will only deteriorate with time. Thus, we can conclude that there is sufficient disease signature in the blood of PD patients by year 2 to robustly identify PD associated genes.

An improvement in accuracy in the models which only use genomic modalities to generate patient networks is not seen in supplementary figure 4. As mentioned, PD is a progressive disorder, thus it would be expected that a stronger molecular signal would be found at later time points. Given that our models reach significance at later time points, it suggests that there is in fact a stronger signal in the genomics but, the similarity between patients based on the MDS-UPDRS embeddings is crucial to overcome the heterogeneity in the cohort.

Table 2 shows a subset of genes and pathways which had the largest significant contribution to prediction. Many of the genes and pathways identified have external evidence for association with PD. For example, *MAPK1* and *MAPK10* belong to the family of mitogen-activated protein kinases and have been implicated in the development of PD (Kim and Choi, 2010). Pathways such as PTEN Regulation (R-HSA-6807070) and Axon Guidance (R-HSA-422475), consists of genes and pathways involved in neuronal cell death and neuronal development and have external evidences linking them to the disease (Ogino et al., 2016; Lin et al., 2009).

Notably, not every gene identified by the model was from the PD associated gene list. For example, the *IER3* gene, which was identified as strongly contributing to predictions in the year 3 model, is an inflammatory response gene with protective factor to apoptosis. The response of this gene has recently been shown to be dysregulated in PD patients, but was not included in the PD gene association set (Barmpa et al., 2024). It’s prominence at this later time point makes intuitive sense due to the blood derived genomic modalities used and, highlights the methodology’s promise for identifying novel molecular targets for PD.

## Discussion

In this study, we show that using LLM-derived clinical text embeddings from the MDS-UPDRS questionnaire to create a clinical patient similarity network, combined with molecular genomics data, enhances patient classification and interpretability in PD. By leveraging contextualised LLM text embeddings, we were able to capture nuanced patterns in questionnaire responses that extend beyond the capabilities of previous methods based on categorical features. Our approach of deriving patient similarity from these patterns provided significant and robust insights into the molecular mechanisms of disease.

Our experiments highlighted key methodological insights. Integrating clinical embeddings derived from patient questionnaires is crucial for capturing phenotypic information, and the manner of encoding plays a significant role in its effectiveness. Previous studies typically treated clinical questionnaire as categorical or numerical data without considering their semantic textual content describing symptoms. While this approach enables straightforward application of conventional machine learning algorithms, it overlooks the qualitative phenotypic information embedded in the text itself. LLM embeddings leverage semantic richness of text and the full context, resulting in improved performance over traditional feature-driven methods.

Our study highlights benefits of using LLM as text embedding model, which provide comparative advantages over conventional encoder-only text architectures. LLMs enable a unified embedding of entire questionnaire sections, overcoming the limitations of smaller context windows of earlier encoder-only models. Furthermore, LLM’s zero-shot capabilities remove the need for supervised training, utilising their extensive pre-trained parameter knowledge. This feature is particularly effective given the standardised nature of questionnaire text, consisting of identical questions and predefined answer options.

We used the baseline time point of the PPMI dataset to derive our optimal model configuration. At baseline, no patient has begun to receive medication and thus, the MDS-UPDRS scores are not being masked by medicative effects. At subsequent time points, we included patients in their ‘OFF’ state, as defined by PPMI. Previous research has identified that medicative effects may still be masking the assessment scores in the PPMI dataset, as the 6-hour window without medication is not sufficiently long for an accurate assessment (Simuni et al., 2016). We found similar evidence supporting this, as the logistic regression model worsens in classification accuracy over time, thus motivating the use of the baseline time point to identify the optimal model configuration. The text embeddings appear to be less affected by this effect, thus further supporting their inclusion in the framework.

We found that these embeddings not only improved classification accuracy but, when integrated with genomic modalities, yielded robust and statistically significant insights into the molecular mechanisms of PD. Our evidence indicates that it is only the combination of clinically insightful text embeddings and genomic modalities containing disease signatures that yield such insights. The clinical embeddings individually are very predictive. This is evident due to the high classification accuracy throughout. Our model learns to classify from the network structure, thus high accuracy is to be expected when classifying HC from PD patients based on PD symptoms. We found that learning from network structure alone will not provide significant insight into the molecular mechanisms guiding the prediction. Our models did not achieve significance in identifying PD associated genes until year 2 of the PPMI dataset. By this time point, it is likely that, on average, the PD population will have progressed in their disease to a point where there are clear markers present in the blood samples from which the genomics were generated. Despite this, there is not a statistically significant increase in the accuracy of models which only used genomics for network generation and prediction. This is likely due to the lingering heterogeneity in the population, resulting in patient similarity relationships which do not capture disease class. This further strengthens the inclusion of both MDS-UPDRS derived text embeddings and genomics for robust and significant insights into PD classification.

The requirement for interpretability of neural network and artificial intelligence models in biomedicine is well known (Esser-Skala and Fortelny, 2023). In this research, we extend the capabilities of two deep learning architectures: PNet and MOGDx. MOGDx is a deep learning framework which combines genomic modalities with patient similarity networks. PNet is a neural network architecture derived from known biological reactions. By combining these two architectures, we have presented a framework which can learn from patient network structures and genomic inputs in an interpretable manner. Further work was done to extend the PNet methodology to include a statistical test to determine enrichment of molecular features identified. This test, alongside classification accuracy, provides confidence that the features identified are robust and significant.

In the PD patient cohort included, we did not differentiate between those with mutations in a known PD gene and those with a sporadic onset of disease (idiopathic). Given the complex pathology of PD, there are likely different mechanisms causing the disease, both between and within these groups, but results in the same final downstream effect (Corti et al., 2011). Our PD cohort is very heterogenous as a result. This makes identifying any molecular signal a difficult task. Our framework, however, has associated a number of genes, notably the *MAPK* genes, 2 years post diagnosis with statistical significance. *MAPK* is a regulating cellular processes in the brain, and is known to be dysregulated in all PD patients (Kim and Choi, 2010). This finding highlights that our framework can identify known molecular signals common to a very heterogenous population without the requirement for patient subtyping.

A promising avenue for future research is to explore scenarios with subtler baseline PD indicators. With comprehensive screening, the PPMI study enrols PD patients who already exhibit distinct diagnostic indicators. This setting limits opportunities to validate the effectiveness of combining clinical embeddings with genomic data for earlier-stage patients. Investigating prodromal cases that later convert to PD diagnoses could provide a robust framework to assess scenarios with less pronounced symptoms.

## Data Availability

Data used in the preparation of this article were obtained [on April, 5th 2022] from the Parkinsons Progression Markers Initiative (PPMI) database (www.ppmi-info.org/access-dataspecimens/download-data), RRID:SCR 006431. For up-to-date information on the study, visit www.ppmi-info.org

## Author contributions statement

C.L. generated all text embeddings, performed comparative analyses and drafted the manuscript. B.R. performed multi-omic analyses, classifications, interpretation and drafted the manuscript. T.I.S supervised the study, revised the manuscript and approved the final version of the manuscript. P.M. and R.E.M. revised the manuscript and approved the final version of the manuscript

## Acknowledgments

This work was supported by the United Kingdom Research and Innovation [grant EP/S02431X/1], UKRI Centre for Doctoral Training in Biomedical AI at the University of Edinburgh, School of Informatics. For the purpose of open access, the author has applied a creative commons’ attribution [CC BY] licence to any author accepted manuscript version arising.

Data used in the preparation of this article were obtained [on April, 5th 2022] from the Parkinson’s Progression Markers Initiative (PPMI) database (www.ppmi-info.org/access-dataspecimens/download-data), RRID:SCR 006431. For up-to-date information on the study, visit www.ppmi-info.org

PPMI – a public-private partnership – is funded by the Michael J. Fox Foundation for Parkinson’s Research and funding partners, including [list the full names of all the PPMI funding partners found on the PPMI Website].

## Supplementary Material

Supplementary Material is available in the accompanying files S1FiguresTables.pdf. The full table of experimental results at baseline is available in S2BaselineResults.csv. A table of experimental results at all time points is available in S3TimePointResults.csv. The full list of significant features at each time point00 is available in S4FeatureSignificance.xlsx.

