## SupplementaryTextFigureTable for "Combining Clinical Embeddings with Multi-Omic Features for Improved Patient Classification and Interpretability in Parkinson’s Disease"

### 1 Description of Omic Processing

#### 1.1 Messenger RNA Expression (mRNA)

Gene counts were accessed from the Parkinson's Progression Marker Initiative (PPMI) repository. Counts were filtered if they had either zero expression or zero variance in all samples. Further processing was performed to removed transcripts with low expression counts as outlined by Love et al. (2014). Finally, sample outliers were removed if they were more than two standard deviations from the mean node connectivity distance.

#### 1.2 DNA Methylation (DNAm)

Raw idat files were accessed from the PPMI repository and normalised using the danet normalisation algorithm via the MethylPipeR package (Cheng, Y et al. (2023)). The 300k CpG sites with the highest variance were selected for analysis to reduce the memory allocation required by the modality.

#### 1.3 Cerebral Spinal Fluid Proteins (CSF)

Projects 151 and 196 containing CSF proteins were accessed from the PPMI repository. Proteins which had zero abundance or zero variance in all samples were removed. Proteins which contained more than 10% missingness in all samples were removed and all other missing values were imputed using mean imputation.

### 2 Supplementary Figures

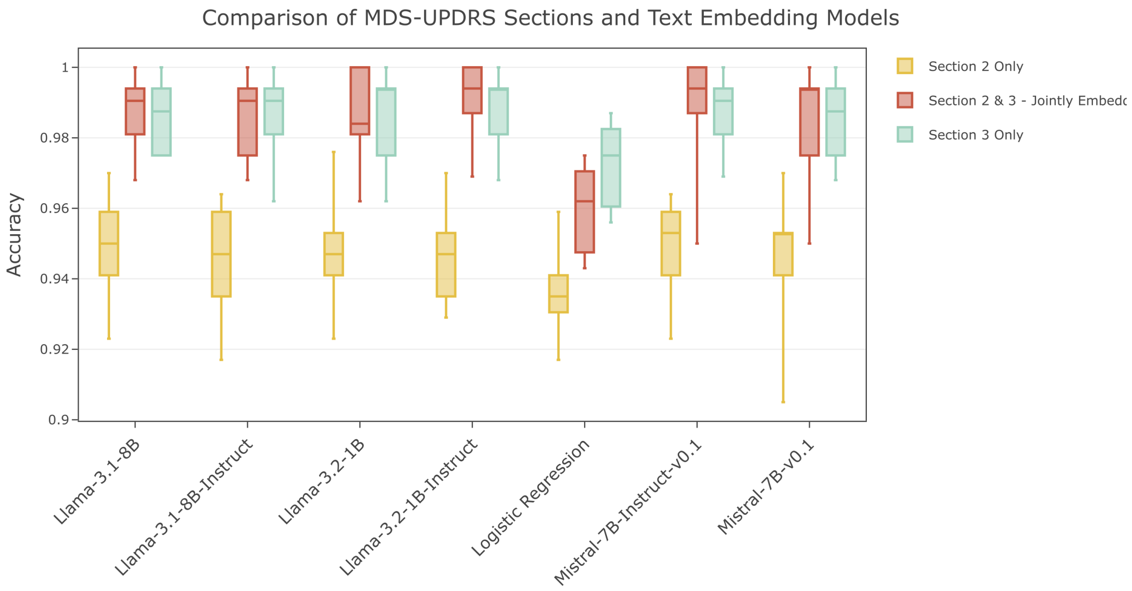


Figure 1: Comparison of MDS-UPDRS Sections and Text Embedding Models used for Patient Similarity Network Generation at Baseline

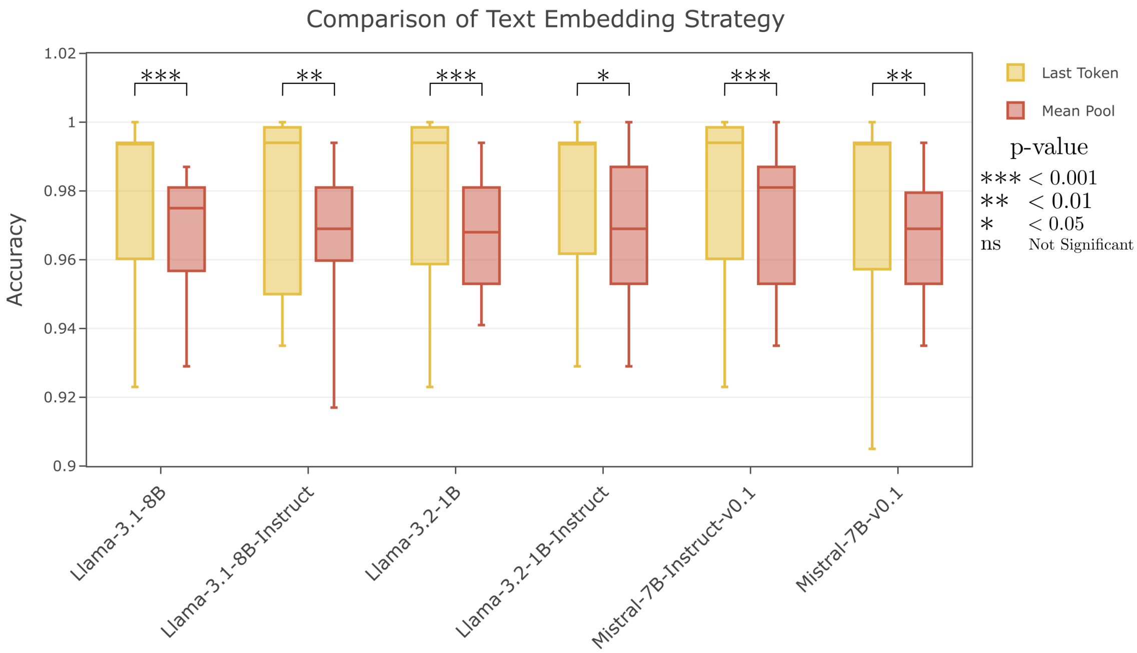


Figure 2: Comparison of Text Embedding Strategies of Mean Pooling and Last Token for Patient Similarity Network Generation at Baseline

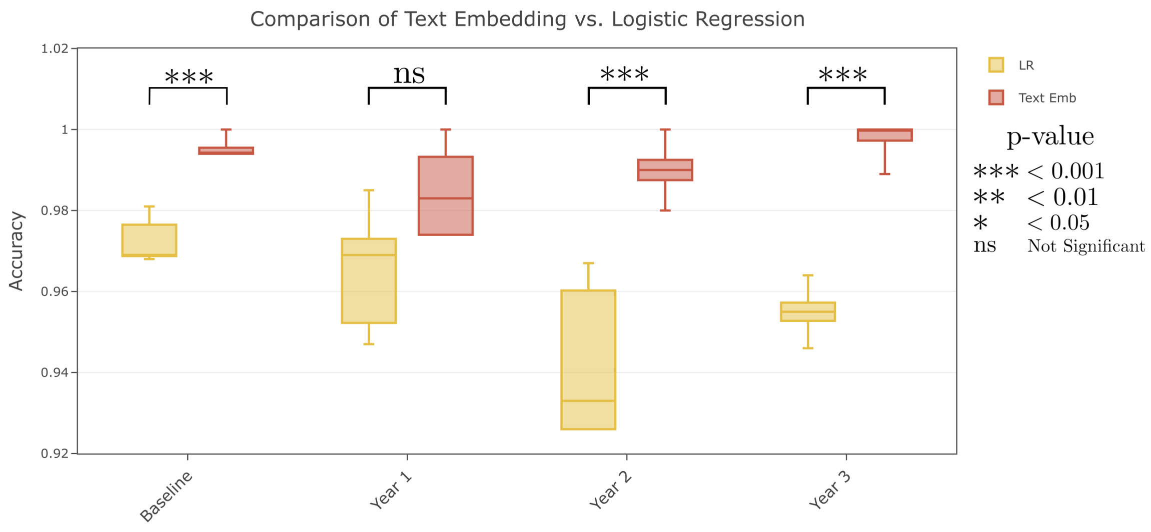


Figure 3: Comparison of Text Embedding vs. Logistic Regression for Patient Similarity Network Generation

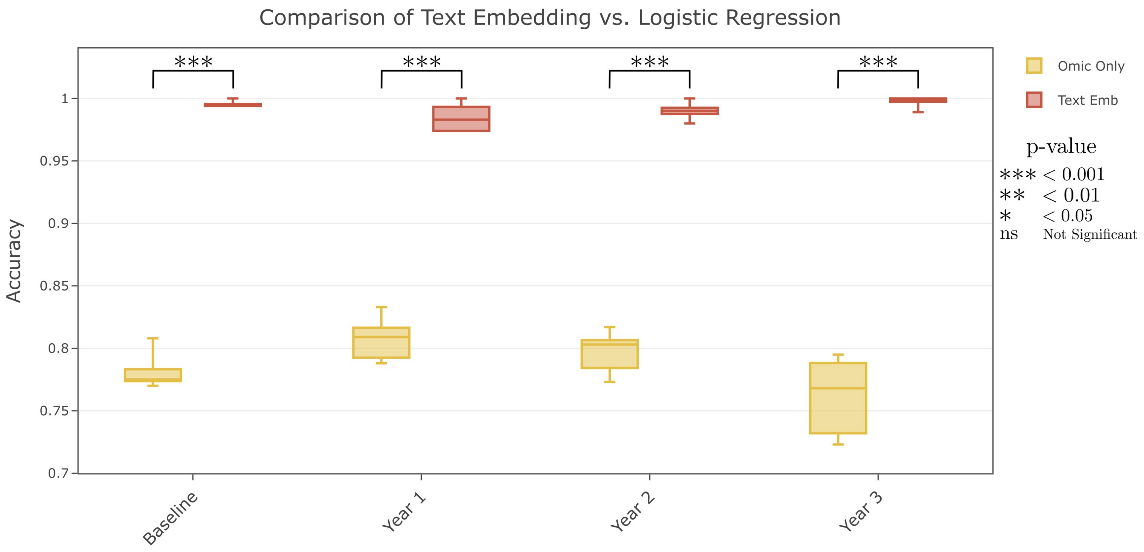


Figure 4: Comparison of Text Embedding vs. Genomic Modalities Only for Patient Similarity Network Generation

The MDS-Unified Parkinson's Disease Rating Scale (MDS-UPDRS) was developed to evaluate various aspects of Parkinson's disease including non-motor and motor experiences of daily living and motor complications. It includes a motor evaluation and characterizes the extent and burden of disease across various populations. Below are the responses of a patient to the MDS-UPDRS questionnaire. Each question is followed by instructions for the patient, examiner, and/or caregiver, as well as the final response.

- - - - - - - - - - - - - - - - - - - - - - - - - - - - - - - - - - - - - - - -

2.1 SPEECH

Over the past week, have you had problems with your speech?
Answer: Normal - Not at all (no problems).

2.2 SALIVA AND DROOLING

Over the past week, have you usually had too much saliva during when you are awake or when you sleep?
Answer: Normal - Not at all (no problems).

2.3 CHEWING AND SWALLOWING

Over the past week, have you usually had problems swallowing pills or eating meals? Do you need your pills cut or crushed or your meals to be made soft, chopped, or blended to avoid choking?
Answer: Normal - No problems.

2.4 EATING TASKS

Over the past week, have you usually had troubles handling your food and using eating utensils? For example, do you have trouble handling finger foods or using forks, knives, spoons, chopsticks?
Answer: Slight - I am slow, but I do not need any help handling my food and have not had food spills while eating.

2.5 DRESSING

Over the past week, have you usually had problems dressing? For example, are you slow or do you need help with buttoning, using zippers, putting on or taking off your clothes or jewelry?
Answer: Slight - I am slow, but I do not need help.

2.6 HYGIENE

Over the past week, have you usually been slow or do you need help with washing, bathing, shaving, brushing teeth, combing your hair, or with other personal hygiene?
Answer: Normal - Not at all (no problems).

2.7 HANDWRITING

Over the past week, have people usually had trouble reading your handwriting?
Answer: Mild - Some words are unclear and difficult to read.

2.8 DOING HOBBIES AND OTHER ACTIVITIES

```
Over the past week, have you usually had trouble doing your hobbies or other
things that you like to do?
Answer:  Normal - Not at all (no problems).

2.9 TURNING IN BED

Over the past week, do you usually have trouble turning over in bed?
Answer:  Normal - Not at all (no problems).

2.10 TREMOR

Over the past week, have you usually had shaking or tremor?
Answer:  Slight - Shaking or tremor occurs but does not cause problems with any
activities.

2.11 GETTING OUT OF BED, A CAR, OR A DEEP CHAIR

Over the past week, have you usually had trouble getting out of bed, a car seat,
or a deep chair?
Answer:  Slight - I am slow or awkward, but I usually can do it on my first try.

2.12 WALKING AND BALANCE

Over the past week, have you usually had problems with balance and walking?
Answer:  Normal - Not at all (no problems).

2.13 FREEZING

Over the past week, on your usual day when walking, do you suddenly stop or freeze
as if your feet are stuck to the floor?
Answer:  Normal - Not at all (no problems).
```

Figure 5: Example input prompt for LLM text embedding from Part 2 of the MDS-UPDRS questionnaire. Input prompts were constructed in the same manner for Part 3 and for a joint input of Parts 2 and 3, where question-response text pairs were concatenated into a single input sequence following a brief overview of the questionnaire.

### 3 Supplementary Tables

| | | mRNA | CSF | DNAm | Total Unique Participants |
|---|---|---|---|---|---|
| Baseline | Healthy Control | 192 | 182 | 86 | 203 |
| | Parkinson's Disease | 704 | 605 | 311 | 816 |
| Year 1 | Healthy Control | 166 | 43 | 85 | 176 |
| | Parkinson's Disease | 440 | 22 | 281 | 482 |
| Year 2 | Healthy Control | 163 | 72 | 82 | 172 |
| | Parkinson's Disease | 439 | 43 | 82 | 487 |
| Year 3 | Healthy Control | 150 | 43 | 81 | 160 |
| | Parkinson's Disease | 333 | 16 | 74 | 401 |

Table 1: Overview of participant availability in PPMI dataset

| | Section 3 Only vs. Section 2 Only | Section 2 & 3 Jointly Embedded vs. Section 3 Only | Section 2 & 3 Jointly Embedded vs. Section 2 Only |
|---|---|---|---|
| Llama-3.2-1B | T-stat = 8.17<br>P-value = 2e-05 | T-stat = -0.44<br>P-value = 0.666 | T-stat= 9.08<br>P-value = 8e-06 |
| Llama-3.2-1B-Instruct | T-stat = 11.3<br>P-value = 1e-06 | T-stat = 2.16<br>P-value = 0.06 | T-stat= 13.74<br>P-value = 2e-07 |
| Llama-3.2-8B | T-stat = 7.4<br>P-value = 4e-05 | T-stat = 0.44<br>P-value = 0.436 | T-stat= 8.29<br>P-value = 2e-05 |
| Llama-3.2-8B-Instruct | T-stat = 10.74<br>P-value = 2e-06 | T-stat = 0<br>P-value = 1 | T-stat= 9.56<br>P-value = 5e-06 |
| Mistral-7B-v0.1 | T-stat = 6.35<br>P-value = 1e-04 | T-stat = 0.5<br>P-value = 0.622 | T-stat= 6.39<br>P-value = 1e-04 |
| Mistral-7B-Instruct-v0.1 | T-stat = 8.76<br>P-value = 1e-05 | T-stat = 0.55<br>P-value = 0.599 | T-stat= 10.84<br>P-value = 2e-06 |
| Logistic Regression | T-stat = 9.80<br>P-value = 6e-04 | T-stat = -51.44<br>P-value = 8e-07 | T-stat= 6.3<br>P-value = 3e-03 |

Table 2: Paired T-test to identify significance between text embedding pooling strategies. For the logistic regression test, sections 2 and 3 were joined by concatenation.
